# Two Data-Driven Approaches to Identifying the Spectrum of Problematic Opioid Use: A Pilot Study within a Chronic Pain Cohort

**DOI:** 10.1101/2021.09.07.21263079

**Authors:** Lori Schirle, Alvin Jeffery, Ali Yaqoob, Sandra Sanchez-Roige, David Samuels

**Affiliations:** Vanderbilt University School of Nursing; Vanderbilt University Department of Biomedical Informatics; Vanderbilt University Data Science Institute; Vanderbilt University Medical Center, Division of Genetic Medicine; University of California Department of Psychiatry; Vanderbilt University School of Medicine, Department of Molecular Physiology and Biophysics, Vanderbilt Genetics Institute

## Abstract

**Background:** Although electronic health records (EHR) have significant potential for the study of opioid use disorders (OUD), detecting OUD in clinical data is challenging. Models using EHR data to predict OUD often rely on case/control classifications focused on extreme opioid use. There is a need to expand this work to characterize the spectrum of problematic opioid use.

**Methods:** Using a large academic medical center database, we developed 2 datadriven methods of OUD detection: (1) a Comorbidity Score developed from a Phenome-Wide Association Study of phenotypes associated with OUD and (2) a Textbased Score using natural language processing to identify OUD-related concepts in clinical notes. We evaluated the performance of both scores against a manual review with correlation coefficients, Wilcoxon rank sum tests, and area-under the receiver operating characteristic curves. Records with the highest Comorbidity and Text-based scores were re-evaluated by manual review to explore discrepancies.

**Results:** Both the Comorbidity and Text-based OUD risk scores were significantly elevated in the patients judged as High Evidence for OUD in the manual review compared to those with No Evidence (p = 1.3E-5 and 1.3E-6, respectively). The risk scores were positively correlated with each other (rho = 0.52, p < 0.001). AUCs for the Comorbidity and Text-based scores were high (0.79 and 0.76, respectively). Follow-up manual review of discrepant findings revealed strengths of data-driven methods over manual review, and opportunities for improvement in risk assessment.

**Conclusion:** Risk scores comprising comorbidities and text offer differing but synergistic insights into characterizing problematic opioid use. This pilot project establishes a foundation for more robust work in the future.

## 1. INTRODUCTION

Despite aggressive increases in opioid epidemic research funding [1], U.S. opioid overdose deaths continue to rise [2,3]. Retrospective observational studies are valuable research tools for examining epidemiology, disease progression, and treatment effectiveness [4], however, their use is hampered in opioid use disorder (OUD) research due to difficulties in OUD detection in Electronic Health Records (EHR) data. Providers are often reluctant to document concerns about opioid use in health records due to the stigmatizing nature of diagnoses, potential difficulties in future pain management, fear of misclassification, and poorly defined diagnostic criteria [5-10]. Therefore, standard approaches for identifying cases in EHR data, such as International Classification of Diseases (ICD) codes or problem lists, insufficiently capture OUD [11-13].

Several existing methods have utilized EHR data for OUD prediction [14-20]. Some models identify OUD cohorts using ICD codes [16-17], but this approach likely underrepresents problematic opioid use [13]. Other models used unstructured clinical notes text [14,15,18,19]. Although useful, both methods characterize problematic opioid use in a binary fashion, missing nuanced problematic opioid use that occurs not in a *present/absent* dichotomy but on a *continuum of severity*.

Other studies employ models that do capture the continuum of problematic opioid use outside of EHR data. For example, one study used machine learning methods to produce a continuous measure of OUD risk in Medicare data purchased through the Centers for Medicare and Medicaid (CMS) [22]. These studies provide indispensable insight into the continuum of problematic opioid use, but come at the expense of poor reproducibility in most EHR systems [13,20-22].

To overcome this limitation, a recent study employed machine learning approaches to produce a continuous measure of problematic opioid use risk using readily accessible inpatient EHR data [23]. Our study expands this foundational work by employing two data-driven methods to assess the continuum of problematic opioid use using different data sources (ICD codes and clinical notes) in a large sample of readily available EHR data comprising all encounters for chronic pain patients. The first method uses a phenome wide association study (PheWAS) of phenotypes significantly associated with OUD ICD codes to produce an OUD comorbidity risk score. The second method uses natural language processing (NLP) to produce a text-based score to identify OUD-related concepts in available clinical notes. Methods to detect the continuum of problematic opioid use in readily available EHR data would significantly enhance the ability to conduct retrospective opioid research critical to improving OUD detection and treatment.

The objective of this study was to evaluate whether combining data-driven OUD comorbidities and EHR text could serve as a new framework to identify a continuum of problematic opioid use.

## 2. Material and Methods

### 2.1 Overall Procedure and Cohort Selection

Data from Vanderbilt University Medical Center’s de-identified biorepository, BioVU, linked to over 20 years of clinical records [24], was extracted between 8/2018 through 6/2021. We developed the ICD-based OUD comorbidity score and text-based Concept Unique Identifier (CUI) score in independent BioVU participant subgroups. To develop the text-based score and evaluate final performance, we chose individuals with a diagnosis of chronic pain due to a higher incidence of opioid use and OUD than in general populations [25]. We evaluated our methods against gold-standard manual review in a holdout test set (Figure 1).

**Figure 1.**
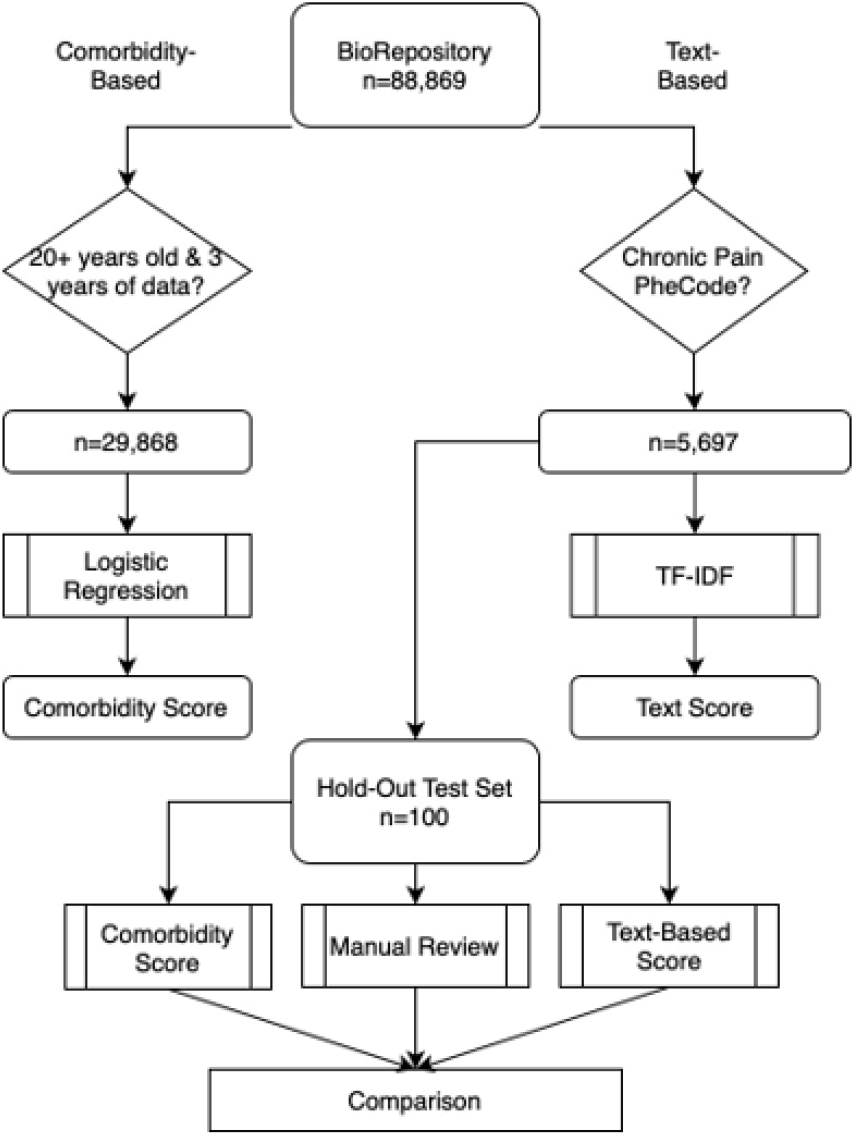
Flow Diagram for Sample Selection of Comorbidity-based and Text-based Problematic Opioid Use Risk Scores

This study follows Strengthening the Reporting of Observational Studies in Epidemiology (STROBE) reporting guidelines. We acquired institutional IRB approval.

### 2.2 OUD Comorbidity Score Development

#### 2.2.1 OUD PheWAS Procedure

For the OUD PheWAS, we used a cohort of Caucasian BioVU participants (N=29,868), as this available dataset was created for genetics analyses, independent of analyses reported here. We required a minimum age at end of the medical record of 20 years, and minimum 3-year length of record [26,27]. Minimum length of record is a common PheWAS practice to improve data depth for each individual [28,29]. This cohort was 42% male, with average record length of 12 years (IQR 7.5 – 15.4 years) and average age at end of medical record of 59 years.

OUD was defined by presence of relevant ICD9 or ICD10 codes (Supplemental Table 1). Individuals with at least one OUD ICD code were classified as a PheWAS OUD case. In this cohort, the OUD rate was 2.1%. PheWAS phenotype categories were defined through presence of ICD codes as defined in Wei et al. [30]. Phenotype categories were tested for association with OUD by logistic regression adjusting for sex, age at final record, record duration, and the first 5 genetic principal components to adjust for population substructure. We tested 1,356 PheWAS categories in separate logistic models. All calculations were carried out in R version 3.6.3. Table 1 lists the top 24 association phenotypes, sorted by *p*-value.

**Table 1.** PheWAS Results for Association with OUD

| Phenotype Name | phecode | Beta | Standard Error | p value | Used in model |
| --- | --- | --- | --- | --- | --- |
| Substance addiction and disorders | phe_316 | 5.49 | 0.14 | < 1E-285 | No |
| Chronic pain | phe_338.2 | 2.952 | 0.082 | 3.64E-285 | Yes |
| Suicidal ideation or attempt | phe_297 | 4.06 | 0.14 | 5.29E-185 | Yes |
| Alcohol-related disorders | phe_317 | 4.49 | 0.16 | 2.27E-183 | No |
| Bipolar | phe_296.1 | 3.57 | 0.12 | 3.35E-182 | Yes |
| Suicidal ideation | phe_297.1 | 4.15 | 0.14 | 7.48E-182 | Yes |
| Pain | phe_338 | 2.095 | 0.074 | 4.44E-176 | Yes |
| Major depressive disorder | phe_296.22 | 3.26 | 0.12 | 1.16E-171 | Yes |
| Alcoholism | phe_317.1 | 4.42 | 0.16 | 1.52E-162 | No |
| Posttraumatic stress disorder | phe_300.9 | 3.80 | 0.14 | 4.35E-159 | Yes |
| Tobacco use disorder | phe_318 | 3.66 | 0.14 | 1.06E-145 | No |
| Personality disorders | phe_301 | 4.11 | 0.16 | 2.71E-139 | Yes |
| Mood disorders | phe_296 | 2.64 | 0.11 | 1.16E-135 | Yes |
| Depression | phe_296.2 | 2.63 | 0.11 | 9.97E-131 | Yes |
| Chronic pain syndrome | phe_355.1 | 2.62 | 0.11 | 2.81E-126 | Yes |
| Anxiety disorders | phe_300 | 2.58 | 0.11 | 2.10E-125 | Yes |
| Anxiety disorder | phe_300.1 | 2.56 | 0.11 | 1.61E-118 | Yes |
| Somatoform disorder | phe_303.4 | 4.12 | 0.18 | 7.63E-117 | Yes |
| Acute pain | phe_338.1 | 1.88 | 0.082 | 3.27E-116 | Yes |
| Dysthymic disorder | phe_300.4 | 3.00 | 0.13 | 3.70E-114 | Yes |
| Psychogenic and somatoform disorders | phe_303 | 3.60 | 0.16 | 4.38E-112 | Yes |
| Suicide or self-inflicted injury | phe_297.2 | 4.30 | 0.19 | 6.74E-109 | Yes |
| Agoraphobia, social phobia, and panic disorder | phe_300.12 | 3.29 | 0.16 | 4.46E-107 | Yes |
| Antisocial/borderline personality disorder | phe_301.2 | 4.33 | 0.20 | 2.00E-105 | Yes |

#### 2.2.2 OUD Comorbidity Score Development

The phenotype most significantly associated with OUD was “Substance Addiction and Disorders”, a broad category including the ICD codes used to define OUD. Since our intention was to define a comorbidity score for OUD, we excluded these phenotype categories and all other substance use disorder categories (Table 1). The remaining top 20 associated phenotypes were used to define the comorbidity score. Interestingly, all phenotypes in this score were either pain or mental illness phenotypes.

All ICD9 and ICD10 codes mapping to the 20 PheWas codes in Table 1 were extracted from all subjects in BioVU. Each person was classified as a case or control for these 20 phenotypes. The comorbidity score was calculated as a weighted linear sum over these 20 phenotypes using the beta values for the PheWAS as weights. The maximum comorbidity score in this cohort (51.509) was used to normalize the comorbidity scores to range from 0 to 1.

To test reproducibility and transferability of the PheWAS results (Table 1) to other independent cohorts and other racial groups, we repeated the association of these phenotypes, defined by lists of ICD codes (Supplemental table 1), in two additional cohorts. First, we repeated association tests in an independent cohort of 13,508 Caucasians over age 20 at the end of their medical record and with at least 3 years of medical record. All 20 tested phenotype associations with OUD replicated in this additional cohort with significant associations (minimum *p-*value was 1.4E-10) and effect sizes in the same direction as the initial cohort. To test transferability of the associations to a non-Caucasian population we repeated the association analysis for these 20 phenotypes with OUD in a set of 8,159 African American patients (with minimum 3 years of record length and age 20 years at end of medical record). All 20 associations replicated with the largest *p*-value being 2.0E-12. Hispanic and Asian populations in BioVU were not large enough to carry out transferability tests. Beta values from the association, which are the basis for comorbidity scores, correlated between Caucasian and African American replication cohorts with an r = 0.67, *p* = 0.0011 (Supplemental Figure 1, Supplementary Table 2).

### 2.3 Text-based Score Development

#### 2.3.1 Pre-Processing

We extracted clinical notes from the time period of 30 days before the patient’s first ICD-9 code related to chronic pain (i.e., 338.2, 338.21, 338.22, 338.28, 338.29) through 30 days after the last ICD-9 code related to chronic pain. We excluded patients with only 1 ICD-9 code related to chronic pain. For computational feasibility, we restricted notes based on Observational Medical Outcomes Partnership (OMOP) note type identifiers 44814645 (“Note”) and 44814640 (“Outpatient Note”) and further restricted to those notes containing words related to variations of: *pain, opioid*, expansions of *narcot-*, and expansions of *addict-*. We processed the resulting 308,264 notes using ScispaCy, which is an open-source natural language processing algorithm that modifies routine natural language processing to accommodate biomedical text [31]. We used ScispaCy for sentence detection, abbreviation expansion, named-entity recognition, and negation detection [32,33].

Following recognition of named entities, we used ScispaCy’s EntityLinker component to map entities (i.e., words and phrases) to the Unified Medical Language System (UMLS) standardized vocabulary’s concept unique identifiers (CUIs). As an example, imagine a brief clinical note: “Patient presents for acute pain in R knee. No history of opioid abuse. Prescribing oxycodone.” ScispaCy would separate this note into 7 named entities that map to CUIs. The resulting representation would be: C0030705 (patients), C0184567 (acute onset pain), C0230431 (structure of right knee), C0332122 (no history of-negated) C0029095 (opioid abuse-negated), C0278329 (prescribed), and C0030049 (oxycodone).

#### 2.3.2 Conversion to Numerical Features

In addition to routine stop-words (e.g., *it, the, and*), we removed 17 ambiguously mapped concepts (e.g., the word “met” in the frequent context of “goals met” was mapped to C0025646 for “methionine”) (Supplemental Table 3). To represent the relative importance of a concept for a patient’s corpus of notes, we calculated Term Frequency-Inverse Document Frequency (TF-IDF) scores for each non-negated CUI for each patient [28,29].

#### 2.3.3 Salient Concept Identification and Score Development

We explored CUIs with the 50 highest TF-IDF values from both patients labeled as cases from ICD codes (Supplemental Table 1), and those labeled as controls. Subject matter experts (L.S., a nurse anesthetist and opioid researcher, and S.S-R., a substance use disorder and genetics researcher) compared top CUIs found only in cases versus top CUIs found only in controls. Table 2 lists top-scoring CUIs for cases, along with whether subject matter experts identified the concept as valid. Top CUI scores included terms such as ‘methadone’ and ‘Suboxone’. For each CUI identified as valid, we added 1 point when a patient’s TF-IDF value for that CUI was larger than the mean TF-IDF value across all patients. We performed a similar process for CUI controls (Supplemental Table 4). However, the inclusion of control data increased noise, and score performance decreased considerably. Therefore, we removed control data from the scores.

**Table 2.**
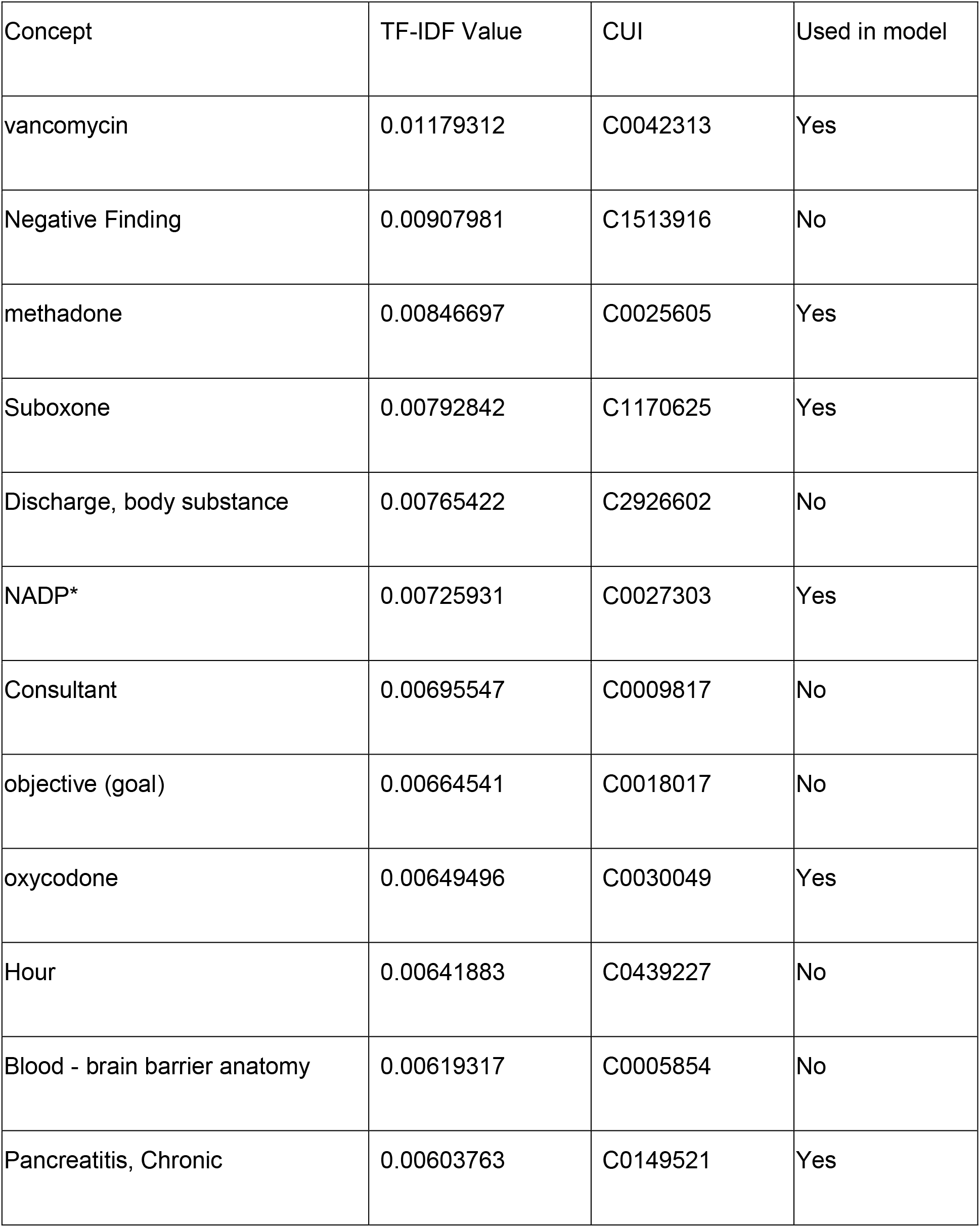
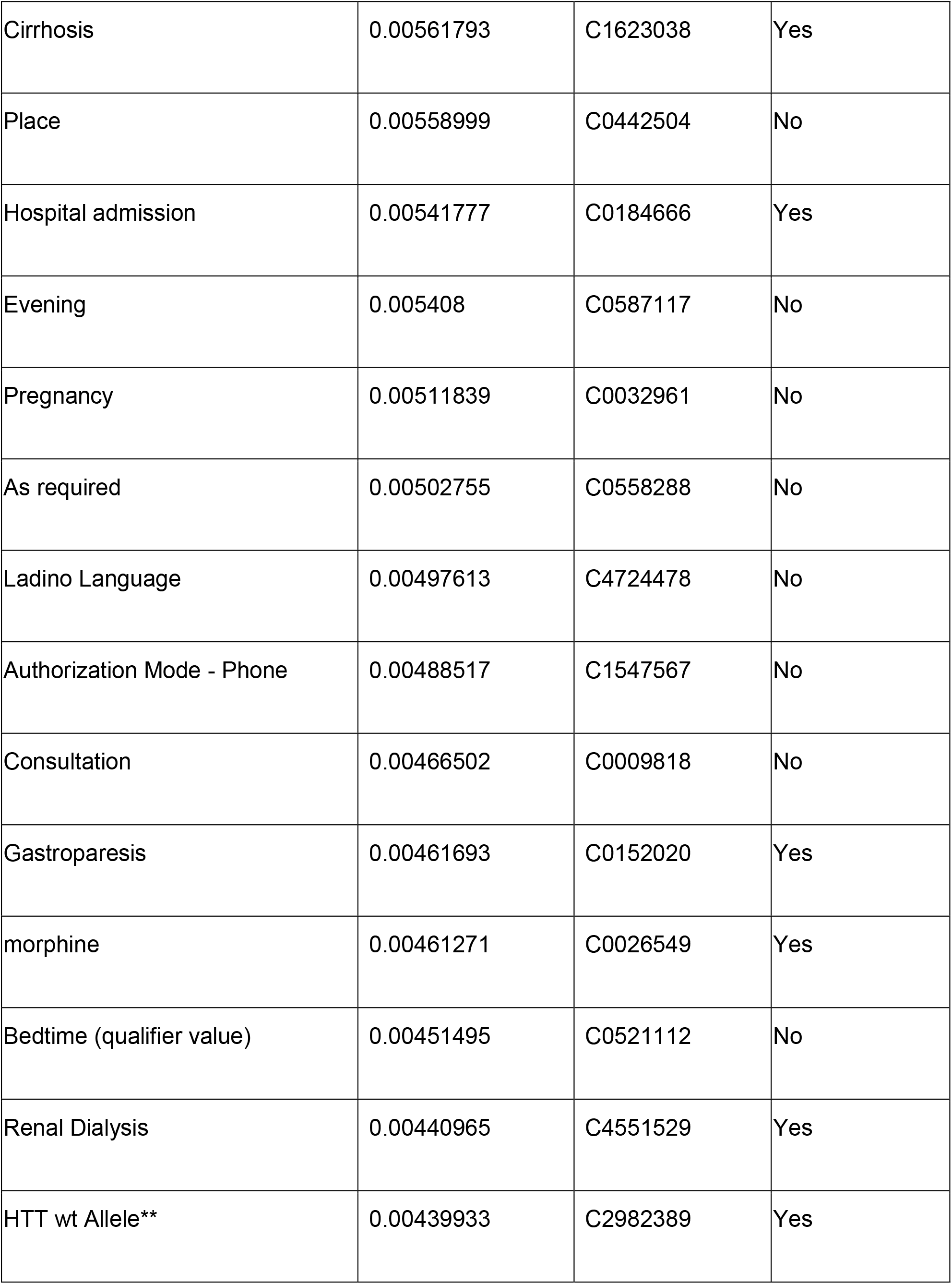
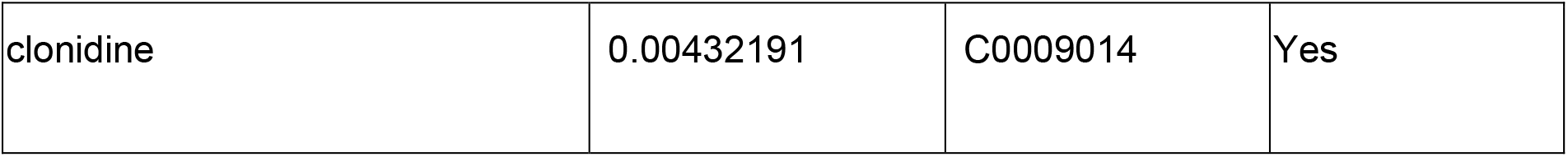
Highest Ranking TF-IDF Values for CUIs Found Only in Cases, Along with Subject Matter Expert Recommendation for Inclusion

### 2.4 Manual Medical Record Review Procedure

To determine evidence of problematic opioid use, a randomly-selected subset of 100 patients from a holdout set of the chronic pain cohort underwent manual record review for comparison with data-driven methods. One record did not contain sufficient data to calculate a text-based score and was excluded from manual review. We reviewed records using a keyword template developed from keywords in the Diagnostic and Statistical Manual of Mental Disorders, 5^th^ Ed. (DSM V) criteria for OUD [34], the Addiction Behaviors Checklist [35], and previous studies describing problematic opioid use detection in EHRs [15,19,20]. Periodic interim analyses assessed word performance, and we trimmed duplicate words (i.e. “detox” and “tox screen” to “tox”, and “multiple providers” and “multiple prescribers” to “multiple pr”). Supplemental Table 5 contains final keywords. Two subject matter experts (L.S., S.S-R) independently reviewed EHR data, and classified patients into one of three categories for evidence of problematic opioid use in health records (No, Some, High). See Supplemental Figure 2 for manual review details. Reviewers were blinded to the patient’s text-based and comorbidity scores.

### 2.5 Statistical Analysis

We conducted statistical analyses in R v3.6.3 and in Python 3.8.5. We used Spearman’s *rho* rank correlation coefficient to examine correlations between each scoring system and manual review as well as the correlation between scoring systems. Comparisons of OUD comorbidity scores and text-based risk scores between manual review categories were carried out by one-sided Wilcoxon rank sum test with continuity correction. We used an area under the receiver operating characteristic curve (AUC) to compare the true positive rate with the false positive rate across all thresholds for both scoring systems.

## 3. Results

### 3.1 Manual Medical Record Review

Upon manual review of 99 chronic pain patients, 49.5% were classified as having No Evidence, 32.3% as having Some Evidence, and 17.2% as having High Evidence for OUD, in line with previously recorded OUD prevalence in chronic pain patients [10].

### 3.2 Comparison of Comorbidity Score, Text-based Risk Score, and Manual Review

Comorbidity scores from High Evidence and Some Evidence groups were significantly higher than the No Evidence group (*p* = 1.3 × 10^−5^, *p* = 2.0 × 10^−4^, respectively; Figure 2A). Comorbidity scores between High and Some Evidence groups were also different, with the High Evidence group having higher comorbidity scores (*p* = 0.039). Similar patterns were observed in the text-based scores (Figure 2A).

**Figure 2.**
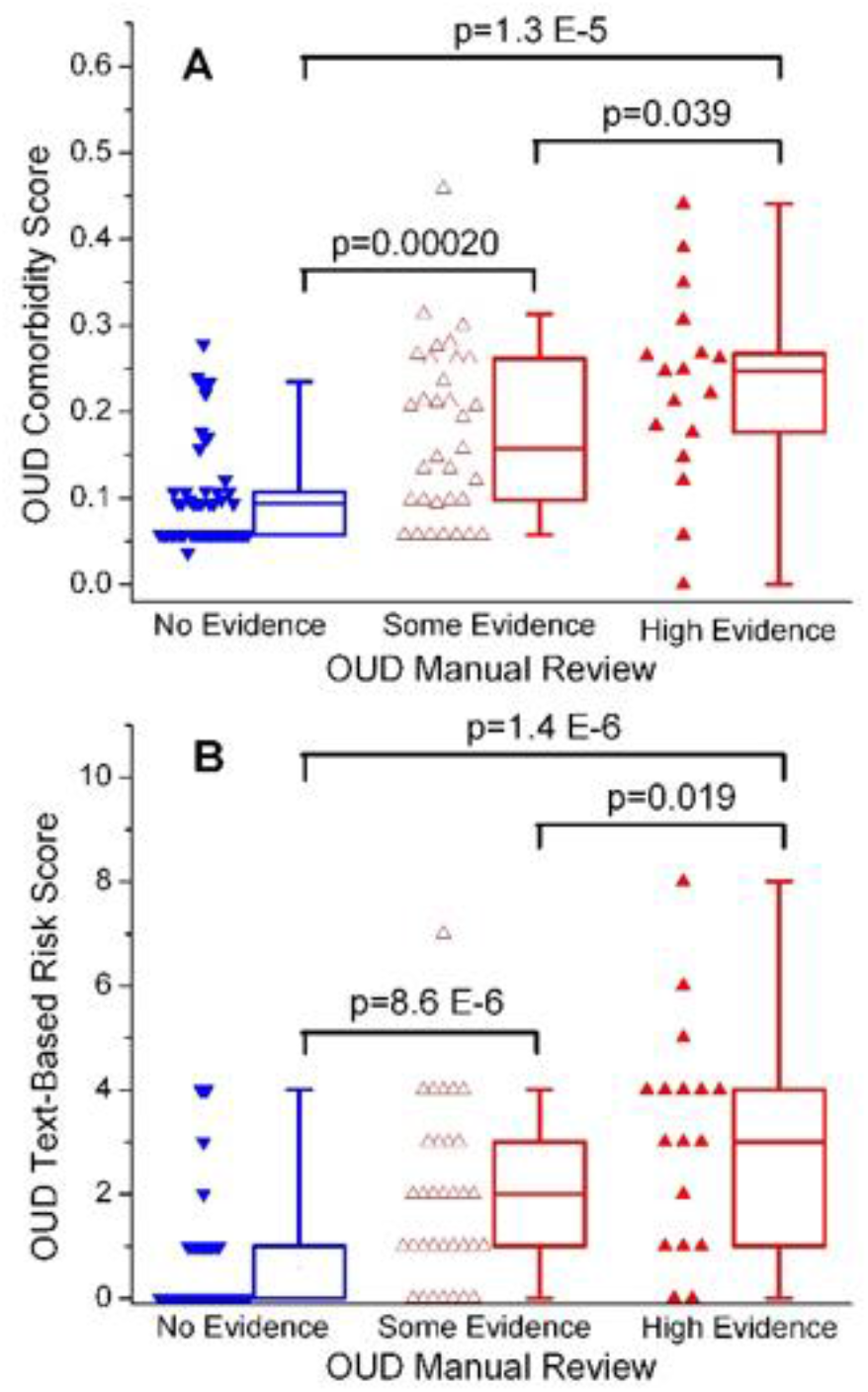
Distribution of Comorbidity and Text-based Risk Scores across Manual Review Categories

The manual review categories for High/Some evidence for OUD were positively correlated with the comorbidity (*rho* = 0.49, *p* < 0.001) and text-based scores (*rho* = 0.56, *p* < 0.001) (Figure 3). Comorbidity and text-based scores were also positively correlated with each other (*rho* = 0.52, *p* < 0.001).

**Figure 3.**
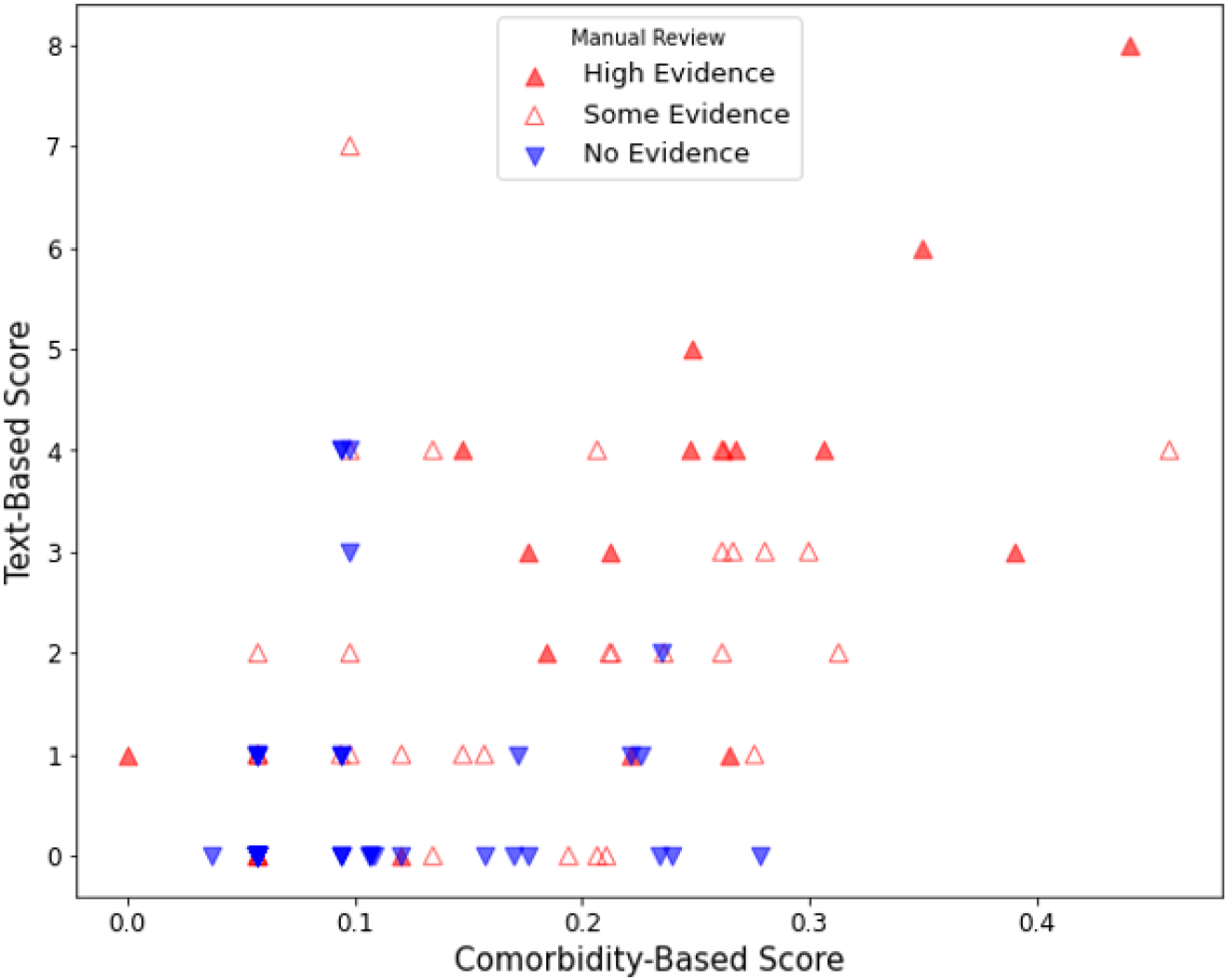
Scatterplot of the Correlation between OUD Comorbidity and Text-based Risk Scores for Problematic Opioid Use Stratified by Manual Review Category

### 3.3 Comparison of the Performance of Comorbidity and Text-based Risk Scores for Problematic Opioid Use

To evaluate the ability of comorbidity and text-based risk scores to detect problematic opioid use, we compared both risk scores to the manual review in the 99 individuals in the hold-out test set (Figure 4). The text-based score achieved an AUC of 0.79, and the comorbidity-score achieved an AUC of 0.76, both indicating moderate-to-high performance.

**Figure 4.**
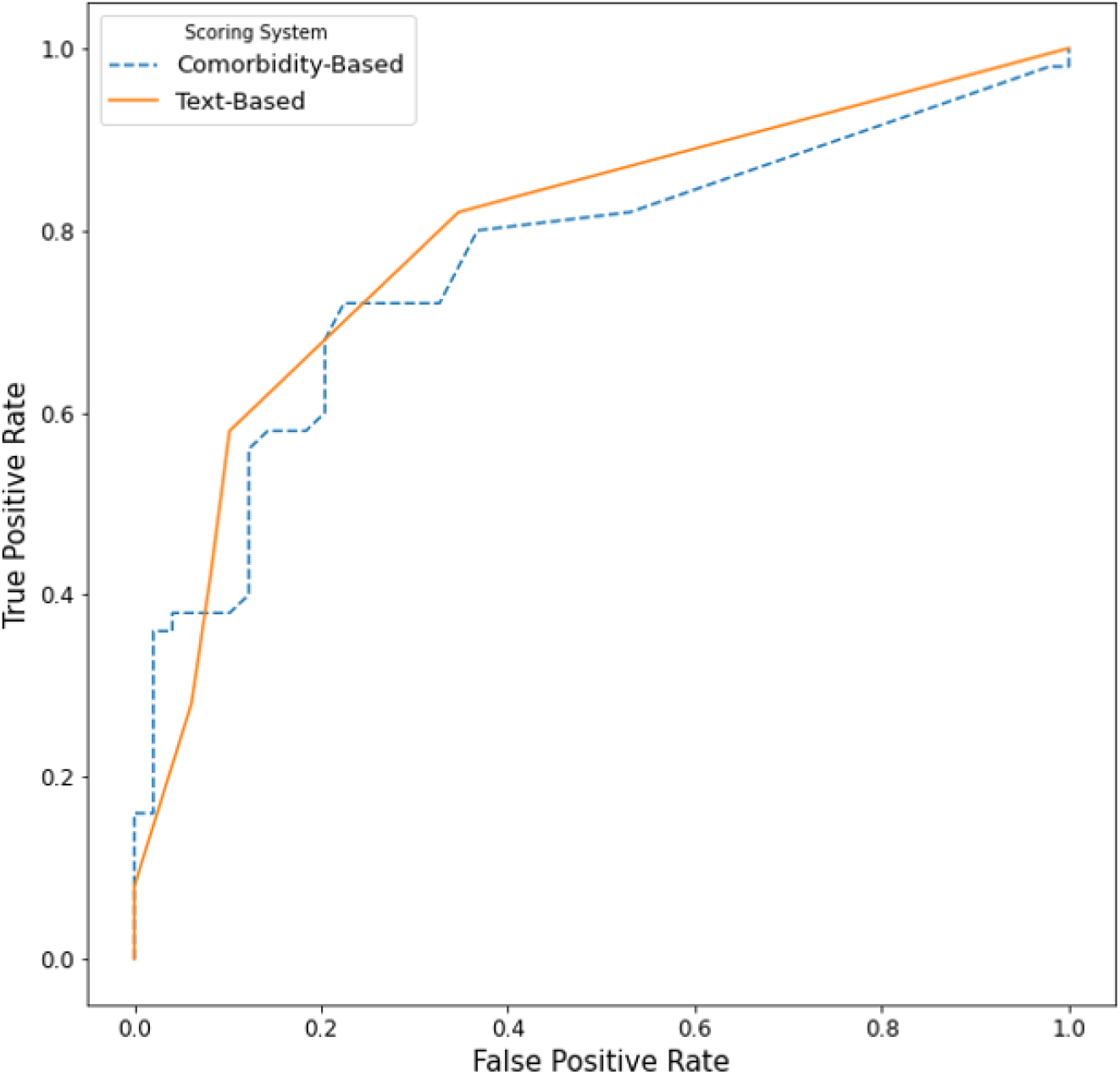
Comparison of the Performance of the Comorbidity and Text-based Risk Scores for Problematic Opioid Use to Manual Review

### 3.4 Post-hoc Manual Review

To investigate concordance between comorbidity scores, text-based scores and manual review results, we considered the individuals scoring in the top quintile of comorbidity scores, and the top quintile of text-based scores, for further follow-up manual review (Figure 5). Table 3 details post-hoc manual review results.

**Figure 5.**
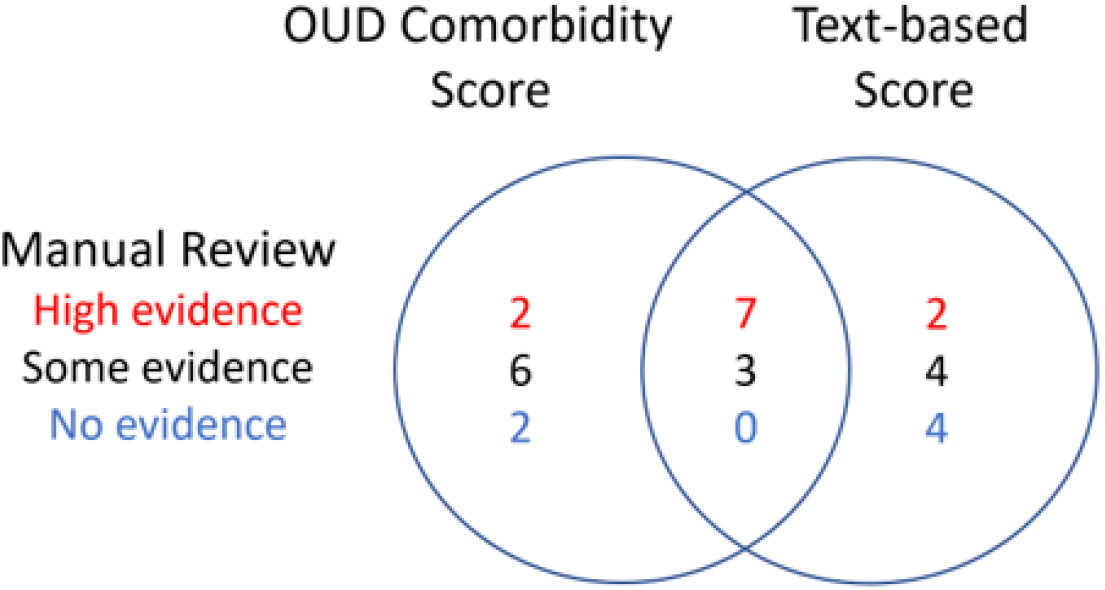
Top Quintiles of OUD Comorbidity and Text-based Scores Stratified by Manual Review Determination of Problematic Opioid Risk

## 4. DISCUSSION

In this pilot study, we developed and tested two data-driven methods to detect OUD in EHR data that helped us characterize the continuum of problematic opioid use. This approach advances existing methods by providing additional benefits surpassing gold standard manual review. In contrast to a manual chart review, our methods increase the objectivity of EHR reviews and could be transferrable to other health care systems with access to ICD codes and clinical notes. Our primary motivation was capturing the continuum of problematic opioid use by assessing indicators of *risk for*, and not *classification of* OUD.

Using these data-driven methods, we identified individuals with high scores who only had limited evidence of OUD in medical records. Notably, these patients were long-term opioid users with indications of potential problematic opioid use but lack the DSM-V signs of compulsive use characteristic of OUD. These individuals may represent a group of chronic pain patients with Complex Persistent Opioid Dependence (CPOD) [7-9]. CPOD, the gray area between opioid dependence and addiction, develops slowly, almost imperceptibly, with long-term opioid exposure [7]. By assessing problematic opioid use risk using a continuous score, these data-driven approaches may identify signs of impending problematic opioid use indiscernible to human clinicians.

An additional advantage of our method is decreased reliance on human data interpretation. For example, two individuals with high data-driven scores were subsequently reassessed by manual review from Some to High Evidence for OUD. In both cases, data critical to (human) review of OUD determination were obscured in records typically not searched in manual review procedures (e.g., phone and intra-provider communications). These data-driven methods rely on agnostic processes not dependent on documented clinician concern for problematic opioid use. Therefore, our method potentially adds to, and expedites, the existing dictionary-based approaches to OUD identification within text [36].

Another major strength of our approach is scalability (ability to evaluate scores quickly over large number of records). Manual review, the gold standard by which OUD EHR detection methods are typically conducted, is extremely labor-intensive, limiting clinical and large-scale research use. Using ICD codes and clinical text, both scores can be adapted to any health system EHR, accommodating regional or system-wide contextual idioms. By using reproducible automated methods in data available in most electronic health systems, these methods have the potential to enhance generalizability.

This work is not without limitations. Due to computational constraints, only text from two note types were included in the analysis; it is possible that valuable information may be captured outside the two note types we explored (e.g. clinical communications). The limited size of the CUI training set may constrain concepts identified and limit transferability across different health systems. Similarly, decisions made in scoring system development could alter their performance. For example, one TF-IDF threshold was set to exclude words found in more than 95% of patients (words found in almost all documents are unlikely to be discriminating). A threshold of 90% or 99% may perform better. Future work to explore threshold influences on scoring system performance is planned. Furthermore, our dependence on a single institution and two types of data could limit our identification of OUD. Additional factors could be used to define a broader cohort (e.g. positive urine screens), but were out of the scope of this project due to limited data available. In addition, we attempted to develop risk scores that captured low or no risk for OUD, but our methods were unable to accurately identify controls. Identifying OUD controls in EHR data is a major limitation of the opioid research field [37]. Interesting insights from the top control CUIs included physical therapy and exercise references (Supplemental Table 4), which will guide future approaches to detect low risk individuals. Lastly, our approach can detect individuals with high probability of opioid misuse. Future studies should examine a broader non-chronic pain population and additional datasets to assess the base rate and dynamics of OUD in other populations, to identify individuals with milder risk, and determine whether these scores can also accurately identify negative cases.

As we acquire larger and more diverse data, we see the scoring systems described here as the first step in developing a clinical decision support tool that could notify clinicians of patients at risk for OUD. We acknowledge concerns that algorithmic approaches to classifying opioid use may lead to medical discrimination [38], the risk for chronic opioid therapy patients to develop problematic opioid use is a prominent concern for clinicians. Identification of these individuals for vigilant monitoring and alternative pain management techniques may be of value in preventing transition from opioid use to OUD.

## Data Availability

Data will be made available upon request

## Notes

### Competing Interest Statement

The authors have declared no competing interest.

### Funding Statement

Jeffery, Alvin: Dr. Jeffery received support for this work from the Agency for Healthcare Research and Quality (AHRQ) and the Patient-Centered Outcomes Research Institute (PCORI) under Award Number K12 HS026395. The content is solely the responsibility of the authors and does not necessarily represent the official views of AHRQ, PCORI, or the United States Government.
The data used for this publication was supported by CTSA award No. UL1 TR002243 from the National Center for Advancing Translational Sciences. Its contents are solely the responsibility of the authors and do not necessarily represent official views of the National Center for Advancing Translational Sciences or the National Institutes of Health.
Computing for this publication was completed at Translational Sciences and the Advanced Computing Center for Research and Education (ACCRE) High-Memory Compute Nodes under Grant No. 1S10OD023680-01 from the National Institutes of Health.
Dr. Sanchez-Roige was supported by funds from the California Tobacco-Related Disease Research Program (TRDRP; Grant Number T29KT0526).
Obtained funding: No unique funding was obtained for this project and no funder input into study design, data acquisition, analysis, or interpretation, or manuscript preparation.

### Author Declarations

This study was deemed exempt by Vanderbilt University IRB

## REFERENCES

1. National Institutes of Health [NIH]. Helping to End Addiction Long-term Initiative [HEAL} 2020 [Available from: https://heal.nih.gov/. Accessed June 21, 2021.

2. Mattson CL, Tanz LJ, Quinn K, Kariisa M, Patel P, Davis NL. Trends and geographic patterns in drug and synthetic opioid overdose deaths — United States, 2013–2019. Vol. 70. 2021. Morbidity and Mortality Weekly Report (MMWR). February 12, 2021. https://www.cdc.gov/nchs/nvss/deaths.htm. Accessed June 21, 2021.

3. Zarefsky M. As COVID-19 surges, AMA sounds alarm on nation’s overdose epidemic. AMA Connect blog. 12/14/2020, 2020. https://www.ama-assn.org/delivering-care/opioids/covid-19-surges-ama-sounds-alarm-nation-s-overdose-epidemic. Accessed 03/03/2021

4. Sessler DI. Big Data–and its contributions to peri-operative medicine. Anaesthesia. Feb 2014;69(2):100–5. doi:10.1111/anae.12537

5. Højsted J, Nielsen PR, Guldstrand SK, Frich L, Sjøgren P. Classification and identification of opioid addiction in chronic pain patients. European journal of pain. 2010;14(10):1014–20.

6. Kovatch M, Feingold D, Elkana O, Lev-Ran S. Evaluation and comparison of tools for diagnosing problematic prescription opioid use among chronic pain patients. International journal of methods in psychiatric research. 2017;26(4):e1542.

7. Manhapra A, Sullivan MD, Ballantyne JC, MacLean RR, Becker WC. Complex persistent opioid dependence with long-term opioids: a gray area that needs definition, better understanding, treatment guidance, and policy changes. J Gen Intern Med. 2020;35(Suppl 3):964–71.

8. Manhapra A, Arias AJ, Ballantyne JC. The conundrum of opioid tapering in long-term opioid therapy for chronic pain: A commentary. Substance Abuse. 2018;39(2):152–61.

9. Ballantyne JC, Sullivan MD, Kolodny A. Opioid dependence vs addiction: a distinction without a difference? Archives of Internal Medicine. 2012;172(17):1342–3.

10. Campbell G, Bruno R, Lintzeris N, Cohen M, Nielsen S, Hall W, et al. Defining problematic pharmaceutical opioid use among people prescribed opioids for chronic noncancer pain: do different measures identify the same patients? Pain. 2016;157(7):1489–98.

11. Dufour R, Joshi AV, Pasquale MK, Schaaf D, Mardekian J, Andrews GA, et al. The prevalence of diagnosed opioid abuse in commercial and Medicare managed care populations. Pain Pract. 2014;14(3):E106–15.

12. Rice JB, White AG, Birnbaum HG, Schiller M, Brown DA, Roland CL. A model to identify patients at risk for prescription opioid abuse, dependence, and misuse. Pain Med. 2012;13(9):1162–73.

13. Sullivan MD, Edlund MJ, Fan MY, DeVries A, Braden JB, Martin BC. Risks for possible and probable opioid misuse among recipients of chronic opioid therapy in commercial and Medicaid insurance plans: The TROUP Study. Pain. 2010;150(2):332–9.

14. Afshar M, Joyce C, Dligach D, Sharma B, Kania R, Xie M, et al. Subtypes in patients with opioid misuse: A prognostic enrichment strategy using electronic health record data in hospitalized patients. PLoS One. 2019;14(7):e0219717.

15. Carrell DS, Cronkite D, Palmer RE, Saunders K, Gross DE, Masters ET, et al. Using natural language processing to identify problem usage of prescription opioids. International journal of medical informatics. 2015;84(12):1057–64.

16. Chartash D, Paek H, Dziura JD, Ross BK, Nogee DP, Boccio E, et al. Identifying opioid use disorder in the emergency department: multi-system electronic health record-based computable phenotype derivation and validation study. JMIR Med Inform. 2019;7(4):e15794.

17. Ellis RJ, Wang Z, Genes N, Ma’ayan A. Predicting opioid dependence from electronic health records with machine learning. BioData Min. 2019;12:3.

18. Hylan TR, Von Korff M, Saunders K, Masters E, Palmer RE, Carrell D, et al. Automated prediction of risk for problem opioid use in a primary care setting. J Pain. 2015;16(4):380–7.

19. Palmer RE, Carrell DS, Cronkite D, Saunders K, Gross DE, Masters E, et al. The prevalence of problem opioid use in patients receiving chronic opioid therapy: computer-assisted review of electronic health record clinical notes. Pain. 2015;156(7):1208–14.

20. Palumbo SA, Adamson KM, Krishnamurthy S, Manoharan S, Beiler D, Seiwell A, et al. Assessment of probable opioid use disorder using electronic health record documentation. JAMA Netw Open. 2020;3(9):e2015909.

21. Che Z, St. Sauver J, Liu H, Liu Y. Deep learning solutions for classifying patients on opioid use. AMIA Annual Symposium Proceedings Archive 2018.

22. Lo-Ciganic W-H, Huang JL, Zhang HH, Weiss JC, Kwoh CK, Donohue JM, et al. Using machine learning to predict risk of incident opioid use disorder among fee-for-service Medicare beneficiaries: A prognostic study. PLOS ONE. 2020;15(7):e0235981.

23. Sharma B, Dligach D, Swope K, Salisbury-Afshar E, Karnik NS, Joyce C, et al. Publicly available machine learning models for identifying opioid misuse from the clinical notes of hospitalized patients. BMC Medical Informatics and Decision Making. 2020;20(1):79.

24. Ritchie MD, Denny JC, Crawford DC, Ramirez AH, Weiner JB, Pulley JM, et al. Robust replication of genotype-phenotype associations across multiple diseases in an electronic medical record. The American Journal of Human Genetics. 2010;86(4):560–572. Doi: 10.1016/j.ajhg.2010.03.003

25. Vowles, KE, McEntee, ML, Julnes, PS, Frohe, T, Ney JP, van der Goes, DN. Rates of opioid misuse, abuse, and addiction in chronic pain: a systematic review and data synthesis. Pain. 2015;156(4):569–576. doi:10.1097/01.j.pain.0000460357.01998.f1

26. Bastarache L, Hughey JJ, Hebbring S, et al. Phenotype risk scores identify patients with unrecognized Mendelian disease patterns. Science. 2018;359(6381):1233.

27. Salvatore M, Beesley LJ, Fritsche LG, et al. Phenotype risk scores (PheRS) for pancreatic cancer using time-stamped electronic health record data: Discovery and validation in two large biobanks. Journal of Biomedical Informatics. 2021;113:103652.

28. Barnado A, Carroll RJ, Casey C, Wheless L, Denny JC, Crofford LJ. Phenome-wide association studies uncover a novel association of increased atrial fibrillation in male patients with systemic lupus erythematosus. Arthritis Care Res (Hoboken). 2018;70(11):1630–1636.

29. Choi L, Carroll RJ, Beck C, et al. Evaluating statistical approaches to leverage large clinical datasets for uncovering therapeutic and adverse medication effects. Bioinformatics. 2018;34(17):2988–2996.

30. Wei WQ, Bastarache LA, Carroll RJ, Marlo JE, Osterman TJ, Gamazon ER, et al. Evaluating phecodes, clinical classification software, and ICD-9-CM codes for phenome-wide association studies in the electronic health record. PLoS One. 2017;12(7):e0175508.

31. ScispaCy. SpaCy models for biomedical text processing. Allenai; 2020.

32. Jurafsky D & Martin JH. Speech and language processing: an introduction to natural language processing, computational linguistics, and speech recognition. 3rd Ed. 2020. https://web.stanford.edu/~jurafsky/slp3/ed3book.pdf

33. Robertson S. Understanding inverse document frequency: on theoretical arguments for IDF. Journal of Documentation. 2004; 60(5):503–520.

34. American Psychiatric Association (APA). Diagnostic and statistical manual of mental disorders, Fifth Edition DSM-5. 5 ed. Washington, DC: American Psychiatric Association; 2013.

35. Wu SM, Compton P, Bolus R, Schieffer B, Pham Q, Baria A, et al. The addiction behaviors checklist: validation of a new clinician-based measure of inappropriate opioid use in chronic pain. J Pain Symptom Manage. 2006;32(4):342–51.

36. Carrell DS, Cronkite D, Palmer RE, Saunders K, Gross DE, Masters ET, et al. Using natural language processing to identify problem usage of prescription opioids. International Journal of Medical Informatics. 2015;84(12):1057–1064.

37. Polimanti R, Walters RK, Johnson EC, et al. Leveraging genome-wide data to investigate differences between opioid use vs. opioid dependence in 41,176 individuals from the Psychiatric Genomics Consortium. Mol Psychiatry. 2020;25(8):1673–1687.

38. Hatoum A, Wendt F, Galimberti M, Polimanti R, Neale B, Kranzler H, et al. Genetic data can lead to medical discrimination: cautionary tale of opioid use disorder. medRxiv; 2020.

