## supplemental figure 1 for "Two Data-Driven Approaches to Identifying the Spectrum of Problematic Opioid Use: A Pilot Study within a Chronic Pain Cohort"

**Supplemental Figure 1. Correlations of the Effect Sizes between African American and Caucasian Top 20 OUD-Associated Comorbidities**

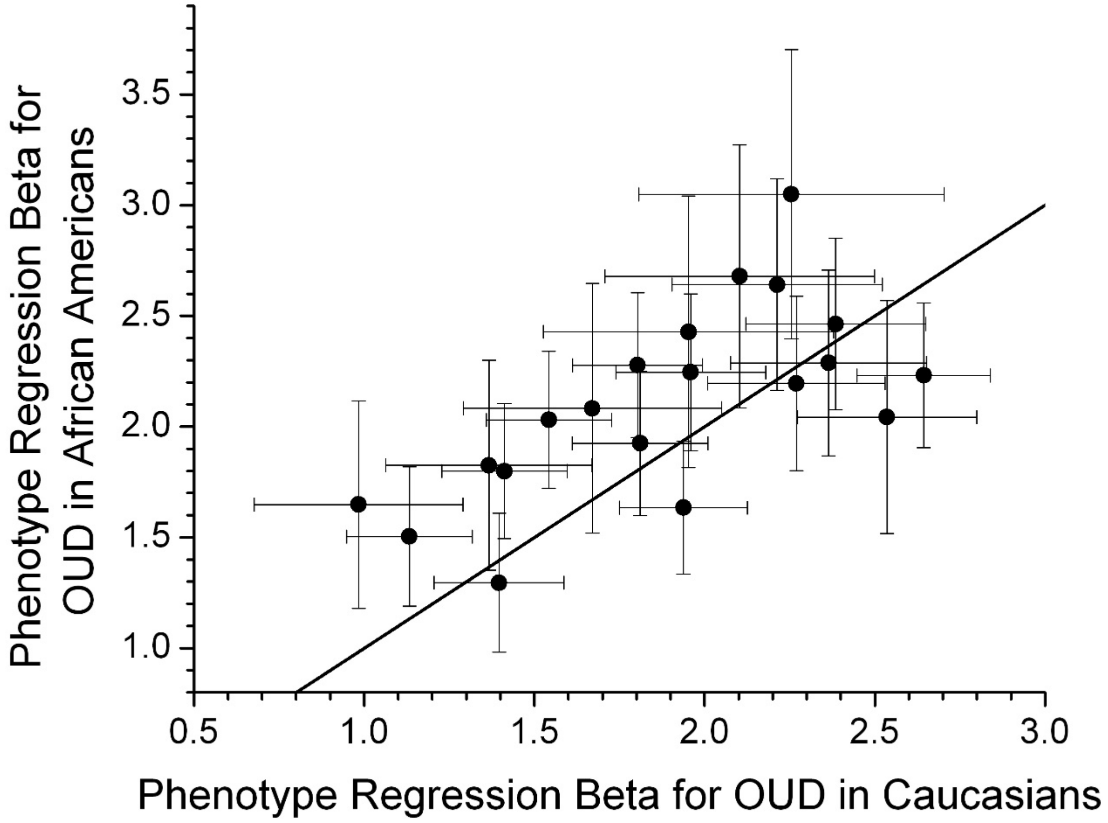


Supplementary Figure 1: Comparison of the phenotype regression beta values for the Caucasian and African American replication cohorts. Error bars are 2 X Standard Error of the regression beta value. The solid line is the line of equal values.
