## Supplementary figures and images for "Two Data-Driven Approaches to Identifying the Spectrum of Problematic Opioid Use: A Pilot Study within a Chronic Pain Cohort"

### supplemental figure 2

**Supplemental Figure 2. Manual Review Process**

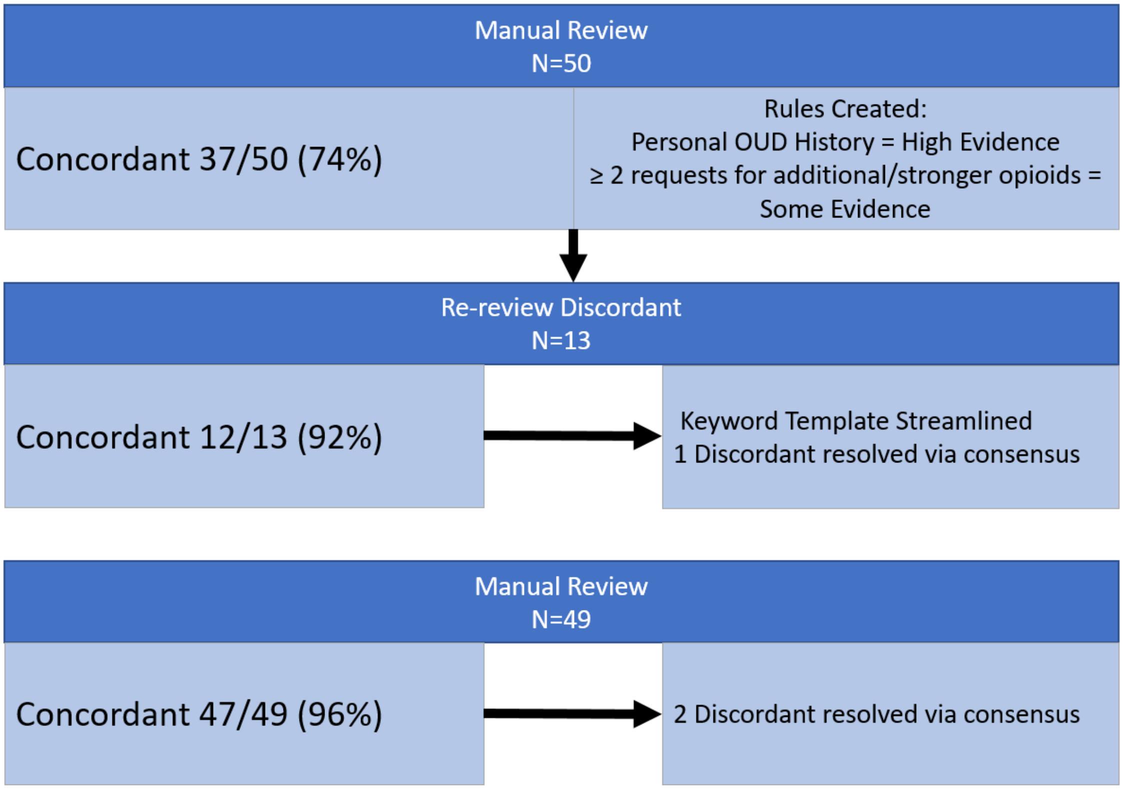
