## supplemental table 1 for "Two Data-Driven Approaches to Identifying the Spectrum of Problematic Opioid Use: A Pilot Study within a Chronic Pain Cohort"

**Supplemental Table 1. ICD9 and ICD10 Codes for Opioid Use Disorder**.

| ICD CODE | CODE_DESC |
|---|---|
| 304.00 | Opioid type dependence, unspecified |
| 304.0 | Opioid type dependence |
| 304.01 | Opioid type dependence, continuous |
| 304.02 | Opioid type dependence, episodic |
| 304.03 | Opioid type dependence, in remission |
| 304.7 | Combinations of opioid type drug with any other drug dependence |
| 304.70 | Combinations of opioid type drug with any other drug dependence, unspecified |
| 304.71 | Combinations of opioid type drug with any other drug dependence, continuous |
| 304.72 | Combinations of opioid type drug with any other drug dependence, episodic |
| 304.73 | Combinations of opioid type drug with any other drug dependence, in remission |
| 305.50 | Opioid abuse, unspecified |
| 305.5 | Opioid abuse |
| 305.51 | Opioid abuse, continuous |
| 305.52 | Opioid abuse, episodic |
| 305.53 | Opioid abuse, in remission |
| F11.10 | Opioid abuse, uncomplicated |
| F11.120 | Opioid abuse with intoxication, uncomplicated |
| F11.121 | Opioid abuse with intoxication delirium |
| F11.122 | Opioid abuse with intoxication with perceptual disturbance |
| F11.129 | Opioid abuse with intoxication, unspecified |
| F11.14 | Opioid abuse with opioid-induced mood disorder |
| F11.159 | Opioid abuse with opioid-induced psychotic disorder, unspecified |
| F11.188 | Opioid abuse with other opioid-induced disorder |
| F11.19 | Opioid abuse with unspecified opioid-induced disorder |
| F11.20 | Opioid dependence, uncomplicated |
| F11.21 | Opioid dependence, in remission |

| | |
|---|---|
| F11.220 | Opioid dependence with intoxication, uncomplicated |
| F11.221 | Opioid dependence with intoxication delirium |
| F11.222 | Opioid dependence with intoxication with perceptual disturbance |
| F11.229 | Opioid dependence with intoxication, unspecified |
| F11.23 | Opioid dependence with withdrawal |
| F11.24 | Opioid dependence with opioid-induced mood disorder |
| F11.251 | Opioid dependence with opioid-induced psychotic disorder with hallucinations |
| F11.259 | Opioid dependence with opioid-induced psychotic disorder, unspecified |
| F11.282 | Opioid dependence with opioid-induced sleep disorder |
| F11.288 | Opioid dependence with other opioid-induced disorder |
| F11.29 | Opioid dependence with unspecified opioid-induced disorder |
| F11.90 | Opioid use, unspecified, uncomplicated |
| F11.920 | Opioid use, unspecified with intoxication, uncomplicated |
| F11.921 | Opioid use, unspecified with intoxication delirium |
| F11.929 | Opioid use, unspecified with intoxication, unspecified |
| F11.93 | Opioid use, unspecified with withdrawal |
| F11.94 | Opioid use, unspecified with opioid-induced mood disorder |
| F11.950 | Opioid use, unspecified with opioid-induced psychotic disorder with delusions |
| F11.951 | Opioid use, unspecified with opioid-induced psychotic disorder with hallucinations |
| F11.959 | Opioid use, unspecified with opioid-induced psychotic disorder, unspecified |
| F11.981 | Opioid use, unspecified with opioid-induced sexual dysfunction |
| F11.982 | Opioid use, unspecified with opioid-induced sleep disorder |
| F11.988 | Opioid use, unspecified with other opioid-induced disorder |
| F11.99 | Opioid use, unspecified with unspecified opioid-induced disorder |
