## supplemental table 2 for "Two Data-Driven Approaches to Identifying the Spectrum of Problematic Opioid Use: A Pilot Study within a Chronic Pain Cohort"

**Supplemental Table 2. Test of Reproducibility and Transferability of OUD Comorbidity Phenotypes.**

| Phenotype Name | phecode | Caucasians (N = 13,508) | | | African Americans (N = 8,159) | | |
|---|---|---|---|---|---|---|---|
| | | Beta | Std Error | p value | Beta | Std Error | p value |
| Chronic pain | phe338.2 | 2.644 | 0.098 | 2.79E-160 | 2.232 | 0.163 | 1.64E-42 |
| Suicidal ideation or attempt | phe297 | 2.270 | 0.131 | 1.13E-67 | 2.195 | 0.197 | 7.37E-29 |
| Bipolar | phe296.1 | 1.959 | 0.110 | 1.24E-70 | 2.245 | 0.177 | 7.97E-37 |
| Suicidal ideation | phe297.1 | 2.364 | 0.144 | 1.15E-60 | 2.288 | 0.210 | 1.22E-27 |
| Pain | phe338 | 1.938 | 0.094 | 5.85E-94 | 1.635 | 0.151 | 1.57E-27 |
| Major depressive disorder | phe296.22 | 1.811 | 0.100 | 1.86E-73 | 1.925 | 0.163 | 3.12E-32 |
| Posttraumatic stress disorder | phe300.9 | 2.385 | 0.132 | 5.05E-73 | 2.463 | 0.194 | 5.57E-37 |
| Personality disorders | phe301 | 2.103 | 0.198 | 2.68E-26 | 2.679 | 0.297 | 2.01E-19 |
| Mood disorders | phe296 | 1.803 | 0.096 | 1.60E-79 | 2.278 | 0.163 | 2.55E-44 |
| Depression | phe296.2 | 1.542 | 0.092 | 6.44E-63 | 2.031 | 0.155 | 2.33E-39 |
| Chronic pain syndrome | phe355.1 | 2.536 | 0.132 | 1.49E-82 | 2.043 | 0.263 | 8.46E-15 |
| Anxiety disorders | phe300 | 1.412 | 0.092 | 3.28E-53 | 1.799 | 0.152 | 3.99E-32 |
| Anxiety disorder | phe300.1 | 1.133 | 0.092 | 1.52E-34 | 1.504 | 0.157 | 1.28E-21 |
| Somatoform disorder | phe303.4 | 1.953 | 0.213 | 5.62E-20 | 2.428 | 0.306 | 2.30E-15 |
| Acute pain | phe338.1 | 1.396 | 0.095 | 1.51E-48 | 1.295 | 0.157 | 1.47E-16 |
| Dysthymic disorder | phe300.4 | 0.984 | 0.153 | 1.39E-10 | 1.648 | 0.234 | 2.01E-12 |
| Psychogenic and somatoform disorders | phe303 | 1.670 | 0.190 | 1.24E-18 | 2.083 | 0.282 | 1.45E-13 |
| Suicide or self-inflicted injury | phe297.2 | 2.213 | 0.154 | 1.14E-46 | 2.641 | 0.239 | 2.01E-28 |
| Agoraphobia, social phobia, and panic disorder | phe300.12 | 1.366 | 0.152 | 2.00E-19 | 1.826 | 0.237 | 1.39E-14 |
| Antisocial/borderline personality disorder | phe301.2 | 2.255 | 0.224 | 7.51E-24 | 3.049 | 0.326 | 8.44E-21 |
