## supplemental table 3 for "Two Data-Driven Approaches to Identifying the Spectrum of Problematic Opioid Use: A Pilot Study within a Chronic Pain Cohort"

**Supplemental Table 3. Rationale for Removing CUIs Based Erroneous Abbreviations**

| CUI | Name | Abbreviation in Source Notes | Assumed Meaning of Abbreviation |
|---|---|---|---|
| C0036644 | Senegal | SN | skilled nursing, student nurse, sensorineural |
| C2828567 | PRSS30P gene | Unknown | Unknown |
| C0032729 | Portugal | PT | patient, physical therapy |
| C0020787 | Idaho | ID | "ID" artifact in de-identified database |
| C2347664 | Preferred term | PT | patient, physical therapy |
| C2348268 | Iodine | I | $1^{st}$ person reference |
| C1422467 | CIA03 gene | Unknown | Unknown |
| C1418208 | OXT gene | OT | occupational therapy |
| C1416596 | KCNK3 gene | 0 – Task | physical/occupational therapy evaluation note artifact |
| C1424863 | CENPJ gene | CPAP | continuous positive airway pressure |
| C1425986 | GBA3 gene | Unknown | found in the basic metabolic panel lab findings |
| C3813711 | TAPBP wt Allele | TPN | total parenteral nutrition |
| C2986618 | LTF wt Allele | LF | date artifact in de-identified database |
| C1332410 | BID gene | BID | twice daily |
| C0025646 | methionine | MET | "met" as in "goals met" |
| C0054696 | carbohydrate-deficient transferrin | CDT | Central Daylight Time |
| C0034802 | epidermal growth factor receptor | EGFR | estimated glomerular filtration rate |
