## supplemental table 4 for "Two Data-Driven Approaches to Identifying the Spectrum of Problematic Opioid Use: A Pilot Study within a Chronic Pain Cohort"

**Supplemental Table 4. Highest Ranking TF-IDF Values for CUIs Found only in Controls, along with Subject Matter Expert Recommendation for Inclusion.**

| Concept | TF-IDF Value | CUI |
|---|---|---|
| Exercise | 0.01234305 | C0015259 |
| Cues | 0.00822126 | C0010439 |
| Knee | 0.00752751 | C0022742 |
| What subject filter - Goals | 0.00726373 | C1546459 |
| Diagnosis:Impression/interpretation of study: Point in time:^Patient:Nominal | 0.00628495 | C0945731 |
| methotrexate | 0.00602848 | C0025677 |
| Migraine Disorders | 0.00589525 | C0149931 |
| Participation Mode - verbal | 0.0058196 | C1548941 |
| microgram | 0.00580524 | C0439211 |
| Structure of left knee | 0.00539489 | C0230432 |
| Physical therapy | 0.00538333 | C0949766 |
| therapeutic aspects | 0.00530983 | C0039798 |
| Structure of right knee region | 0.00530301 | C4281598 |
| Physical therapy exercises | 0.00498423 | C0452240 |
| Upper case Roman letter I | 0.00493458 | C1706104 |
| Facial Hemiatrophy | 0.00463657 | C0015458 |
| Columnar Cell Change of the Breast | 0.00449322 | C1707444 |
| cholecalciferol | 0.00447398 | C0008318 |
| Bilateral | 0.0044581 | C0238767 |

| | | |
|---|---|---|
| Sleep Apnea, Obstructive | 0.00442593 | C0520679 |
| Males | 0.00436086 | C0086582 |
| Absolute | 0.00390596 | C0205344 |
| Structure of right shoulder region | 0.00385283 | C0524468 |
| Money or percentage indicator - Percentage | 0.00373903 | C1561533 |
| APP gene | 0.00373686 | C1364818 |
| Atrial Fibrillation | 0.00370154 | C0004238 |
| vitamin B12 | 0.00369293 | C0042845 |
