## supplemental table 5 for "Two Data-Driven Approaches to Identifying the Spectrum of Problematic Opioid Use: A Pilot Study within a Chronic Pain Cohort"

**Supplemental Table 5. Keyword Template for Standardized Manual Review**

| **Keyword** |
|---|
| opioid |
| opiate |
| pain med |
| narcotic |
| addict(ion) |
| overdose |
| tox (detox or tox screen) |
| UDS |
| quit (drugs, opioids, pain meds, etc) |
| wean (difficulty, failure) |
| illicit |
| aberrant |
| ran/run out |
| early refill |
| calling again (pain med, refill) |
| stolen (prescription, pills) |
| lost (prescription, pills) |
| out of (town, pain med) |
| Multiple pr(escribers, oviders) |
| drug seeking |
| polysubstance |
| on the street |
| contract (opioid) |
| agreement (opioid) |
| legal (difficulties due to opioid use) |
| craving |
| abuse |

| |
|---|
| dependen(t or ce, opioid) |
| Toleran(t or ce, opioid) |
